# Estimated Incidence of Respiratory Hospitalizations Attributable to RSV Infections across Age and Socioeconomic Groups

**DOI:** 10.1101/2022.03.23.22272830

**Authors:** Zhe Zheng, Joshua L. Warren, Eugene D. Shapiro, Virginia E. Pitzer, Daniel M. Weinberger

## Abstract

**Background:** Surveillance for respiratory syncytial virus (RSV) likely captures just a fraction of the burden of disease. Understanding the burden of hospitalizations and disparities between populations can help to inform upcoming RSV vaccine programs and to improve surveillance.

**Methods:** We obtained monthly age-, ZIP code- and cause-specific hospitalizations in New York, New Jersey, and Washington from the US State Inpatient Databases (2005-2014). We estimated the incidence of respiratory hospitalizations attributable to RSV by age and by socioeconomic status using regression models. We compared the estimated incidence and the recorded incidence (based on ICD9-CM) of RSV hospitalizations to estimate the under-recorded rate in the different sub-populations.

**Results:** The estimated annual incidence of respiratory hospitalizations due to RSV was highest among infants <1 year of age with low socioeconomic status, (2700 per 100,000 people, 95% CrI [2600, 2900]) and were considerable in older adults that are ≥65 years of age across socioeconomic status, ranging from 130 to 970 per 100,000 people. The incidence of hospitalization recorded as being due to RSV represented a significant undercount, particularly in adults. Only <5% of the estimated RSV hospitalizations were captured for the older adults that are ≥65 years of age.

**Conclusions:** RSV causes a considerable burden of hospitalization in young children and in older adults in the U.S, with variation by socioeconomic group. Estimates of the incidence of hospitalizations due to RSV in older adults based on the recorded diagnoses likely represent an underestimate.

**Key messages:** *What is already known on this topic?:* Previous study suggested that the incidence of hospitalization for bronchiolitis (often caused by RSV) in infants varies greatly between communities of different socioeconomic levels. However, it is unclear whether these differences are associated with the admission criteria or the risk of RSV infection.

*What this study adds?:* The estimated annual incidence of respiratory hospitalizations due to RSV in infants residing in low socioeconomic areas was about twice as high as the incidence in infants residing in high socioeconomic areas. In older adults, a small fraction of the hospitalizations estimated to be caused by RSV were recorded as such in the inpatient database.

*How this study might affect research, practice or policy?:* With several vaccines and extended half-life monoclonal antibodies against RSV under active development, these estimates can help anticipate the impact of RSV prevention strategies in populations of different demographic characteristics.

## Introduction

Respiratory syncytial virus (RSV) causes a large burden of disease in infants, young children, and the elderly ^1 2^. Although no vaccine against RSV is currently available, multiple vaccines and extended-duration prophylaxis are in development, and preliminary estimates of their efficacy are promising ^3 4^. To accurately assess the potential impact of interventions against RSV, it is essential to evaluate the health burden of RSV in sub-populations with different demographic and socioeconomic characteristics.

Though the epidemic dynamics of RSV vary even within local geographic areas ^5 6^, little is known about how the burden of RSV varies between populations. Previous research has had mixed conclusions about the impact of socioeconomic status (SES) on the risk of RSV infections ^7-9^. While some of the studies found lower rates of RSV infection in higher SES groups, others found no relationship. The incidence of hospitalization for bronchiolitis in infants (often caused by RSV) varies greatly between communities of different socioeconomic levels ^10-12^.

Rates of hospitalizations recorded as being due to RSV infections can also be influenced by testing and coding practices. RSV-associated hospitalizations are likely under-ascertained using laboratory surveillance data. The decision to conduct a diagnostic test for RSV infection and the sensitivity of the tests can influence the completeness of detection. For instance, compared with children, adults are less likely to be tested for RSV, and the diagnostic tests have lower sensitivity in adults ^13 14^.

The goal of this study was to quantify the annual incidence of hospitalizations associated with RSV in different localities and to estimate the degree of under-recording between sub-populations in different localities. To achieve this, we fitted regression models to time series data from comprehensive hospitalization databases from three states in the United States (US) to estimate variations in RSV-attributable incidence of respiratory hospitalizations. Our results can be used to improve RSV surveillance and to inform the potential impacts of different strategies to prevent RSV infections.

## Materials and Methods

### Overview

We used hierarchical Bayesian regression models to estimate the incidence of respiratory hospitalizations attributable to RSV infections in different age and SES groups. The model incorporated monthly data on the number of hospitalizations recorded as due to RSV infections (based on ICD-9-CM) while adjusting for influenza infections, seasonality and underlying temporal trends. The goal of these analyses was to link variations in the number of hospitalizations recorded as due to RSV infections with variations in the number of all-cause respiratory hospitalizations ^15-18^.

### Data sources

Data consisting of individual-level hospital discharges from the states of New York, New Jersey, and Washington from July 2005 to June 2014 were obtained from the State Inpatient Databases of the Healthcare Cost and Utilization Project, maintained by the Agency for Healthcare Research and Quality (purchased through the HCUP Central Distributor) ^19^. Variables included age at admission, ZIP code of residence, the recorded ICD-9-CM-diagnoses (multiple fields),and the month and the year of hospital admission. The outcome in the regression was the number of hospital discharges for respiratory causes by age, month, and SES classification of ZIP code of residence based on median household income at the ZIP-code level. Respiratory hospitalizations were defined as an ICD-9-CM code in the range 460-519 in any of the diagnosis fields (including hospitalizations with diagnoses of pneumonia, bronchitis, and chronic lower respiratory disease (CLRD), etc.). We analyzed hospitalizations in 9 age categories (<1, 1, 2-4, 5-9, 10-19, 20-44, 45-64, 65-84, 85+ years).

Median household income at the ZIP-code level was obtained from the US Census Bureau’s American Community Survey. Our analyses were based on the median household income for 2006-2011. We grouped the ZIP codes into three distinctive SES groups based on the tertile of income. ZIP codes were assigned to the same SES group for the entire period. Population estimates for each ZIP code and age group were obtained from the US Bureau of the Census for the year 2010 ^20^.

The indicator of RSV infections in all age groups was defined as the monthly RSV-specific hospitalizations count (ICD-9-CM code: 079.6, 466.11, 480.1) among children under two years of age in the same SES group. We used RSV infections among children under two years of age as the marker for RSV infections because previous research has shown detection of RSV in children is more accurate than detection in adults ^21 22^. Similarly, to form a more reliable indicator of influenza infections, we aggregated the influenza-specific hospitalizations count (ICD-9-CM code: 487) across age groups in the same SES group.

Time series of monthly count were created for each income level and age group based on the date of hospital admission for either RSV infections or influenza infections and age category. Data cleaning was performed with SAS software, version 9.2 (SAS Institute, Cary, North Carolina). Statistical analyses were performed in R v4.0.2.

### Statistical model

We estimated the age- and SES-specific incidence of respiratory hospitalizations attributable to RSV using hierarchical Bayesian regression models. The model had a negative binomial likelihood and identity link. The hierarchical structure provides advantages compared with fitting these models individually by group. The model pools information across all groups leading to a reduction in uncertainty during parameter estimation while still allowing for differences between groups ^23^. The identity link ensures that each covariate has an additive, rather than a multiplicative, effect on the outcomes of interest ^24 25^. Monthly and yearly dummy variables were used to adjust for seasonality and temporal trends, respectively. Our model used seasonal dummy variables instead of polynomial time trends or sinusoidal curves because the seasonal dummy variable resulted in significantly improved model fit as measured by deviance information criterion (DIC) (Table S1) ^17 18 26^. Details on the model structure can be found in the supplementary document with code available via GitHub (https://github.com/Weinbergerlab/RSVhospitalizations_USA/blob/main/RSV_burden.Rmd).

After we fitted the model, the estimated “true” number of hospitalizations attributable to RSV infections for each time point and stratum was estimated by multiplying posterior samples of the age-specific scaling factor by the count of monthly hospitalizations that were recorded as due to RSV infections among children under 2 years of age in the same SES group. The average annual incidence of hospitalizations attributable to RSV infections was estimated by dividing the sum of the group-specific estimated number of hospitalizations attributable to RSV infections over nine epidemiologic years by the age- and site-specific population. The recording ratios were calculated by dividing the number of recorded ICD-9-CM diagnoses for RSV in each age and SES group by the modeled estimates in the same group. The attributable percent of RSV was calculated by dividing the sum of the estimated number of respiratory hospitalizations attributable to RSV infections by the sum of the model-predicted number of all-cause respiratory hospitalizations in each age and SES group over the entire study period. (Note that model-predicted number of all-cause respiratory hospitalizations and recorded number of all-cause respiratory hospitalizations in each group were almost identical; The model captured 99.98% of the variability in the recorded respiratory hospitalizations in each group.) For each measure, we obtained and summarized samples from the posterior distributions of interest. The incidence was rounded to two significant figures. The recording ratios and attributable percent were rounded to the nearest whole number.

## Results

### Recorded hospitalizations

From July 2005 through June 2014, there were a total of 9,418,390 hospitalizations for respiratory illnesses that contained detailed age and ZIP code information across the three states. Age or ZIP code were missing for 0.1% of the records in the database, and these records were excluded. There were 66,679 hospitalizations recorded as being due to RSV infections among children under two years of age during the study period. State-specific information can be found in the supplementary document (Table S1).

### Estimates of RSV-Associated Respiratory Hospitalizations

The highest estimated incidence of hospitalizations attributable to RSV infections was found in infants <1 year of age (2800 per 100,000 people, 95% CrI [2600, 2900]), followed by children 1-2 years of age (930 per 100,000 people, 95% CrI [830, 1000]) and by adults ≥85 years of age (970 per 100,000 people, 95% CrI [680, 1300]) (**Figure 1**). The estimated hospitalization incidence for RSV infections among children in the ZIP codes from the lowest tertile for SES was almost double that of children in the highest SES ZIP codes (**Figure 1)**. Analyzing the data from each state separately yielded similar results (see Figure S1-S3).

**Figure 1.**
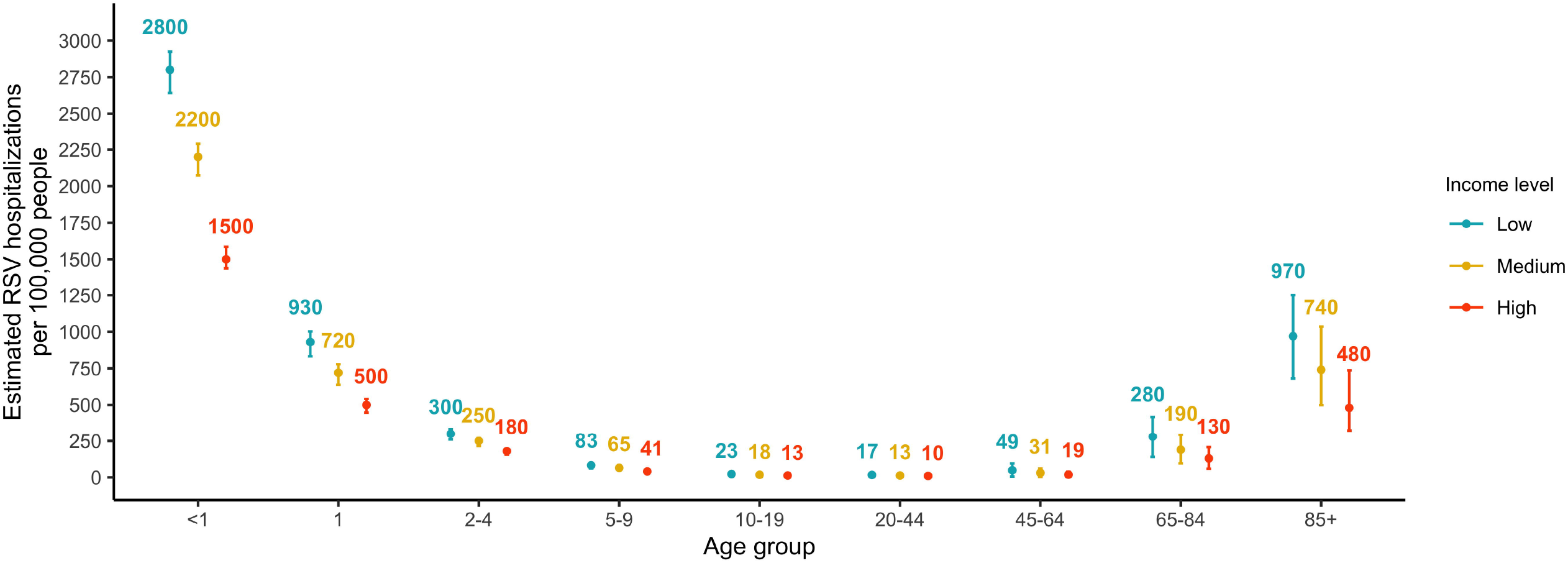
Estimated RSV-attributable respiratory hospitalization rates by age and SES group, July 2005 - June 2014. The color texts show the mean estimates of RSV-attributable respiratory hospitalization rates in each age and SES group. The error bars indicate the 95% credible intervals of the estimated RSV-attributable respiratory hospitalization rates. Color blue, yellow, and orange correspond to the estimates in populations from low, medium, and high SES ZIP codes.

### Gap between recorded and estimated RSV hospitalizations

The estimated proportion of hospitalizations caused by RSV that were recorded as being due to RSV was similar between SES groups and decreased with age (Figure 2). For example, in the highest SES group, 72% (95% CrI: 69%-76%) of estimated RSV hospitalizations among infants under 1 year of age were recorded as being caused by RSV, whereas only 3% (95% CrI: 2%-5%) of hospitalizations estimated to be due to RSV among adults ≥85 years of age were recorded as due to RSV.

**Figure 2.**
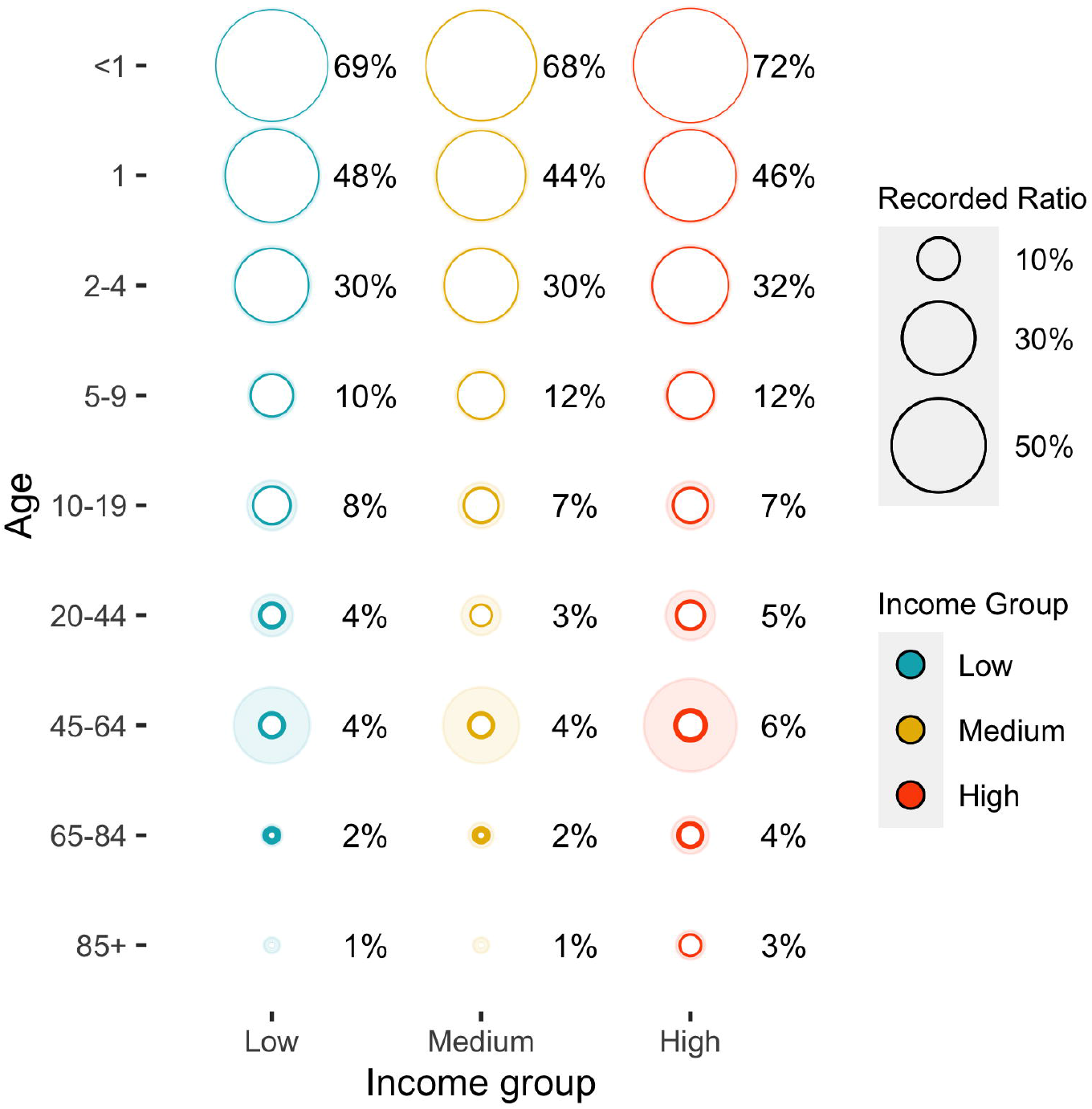
Ratio of the number of hospitalizations recorded as being due to RSV and the number of estimated RSV hospitalizations, by age and SES group, July 2005 - June 2014. The size of the circles represents the estimated proportion of hospitalizations caused by RSV that were recorded. The texts show the mean estimates of the recording ratio in each age and SES group. The shaded areas indicate the 95% credible intervals of the reporting ratio. Color blue, yellow, and orange correspond to the estimates in populations from low, medium, and high SES ZIP codes.

While most of the estimated hospitalizations attributable to RSV infections among adults ≥65 years of age were not recorded as being due to RSV, the rate of recording increased over time (Figure 3). The estimated recording ratio was 1% (95% CrI: 1%-1%) in January (peak time of RSV epidemics) during the 2005-2006 RSV season and increased to 12% (95% CrI: 9%-18%) in January during the 2013-2014 RSV season.

**Figure 3.**
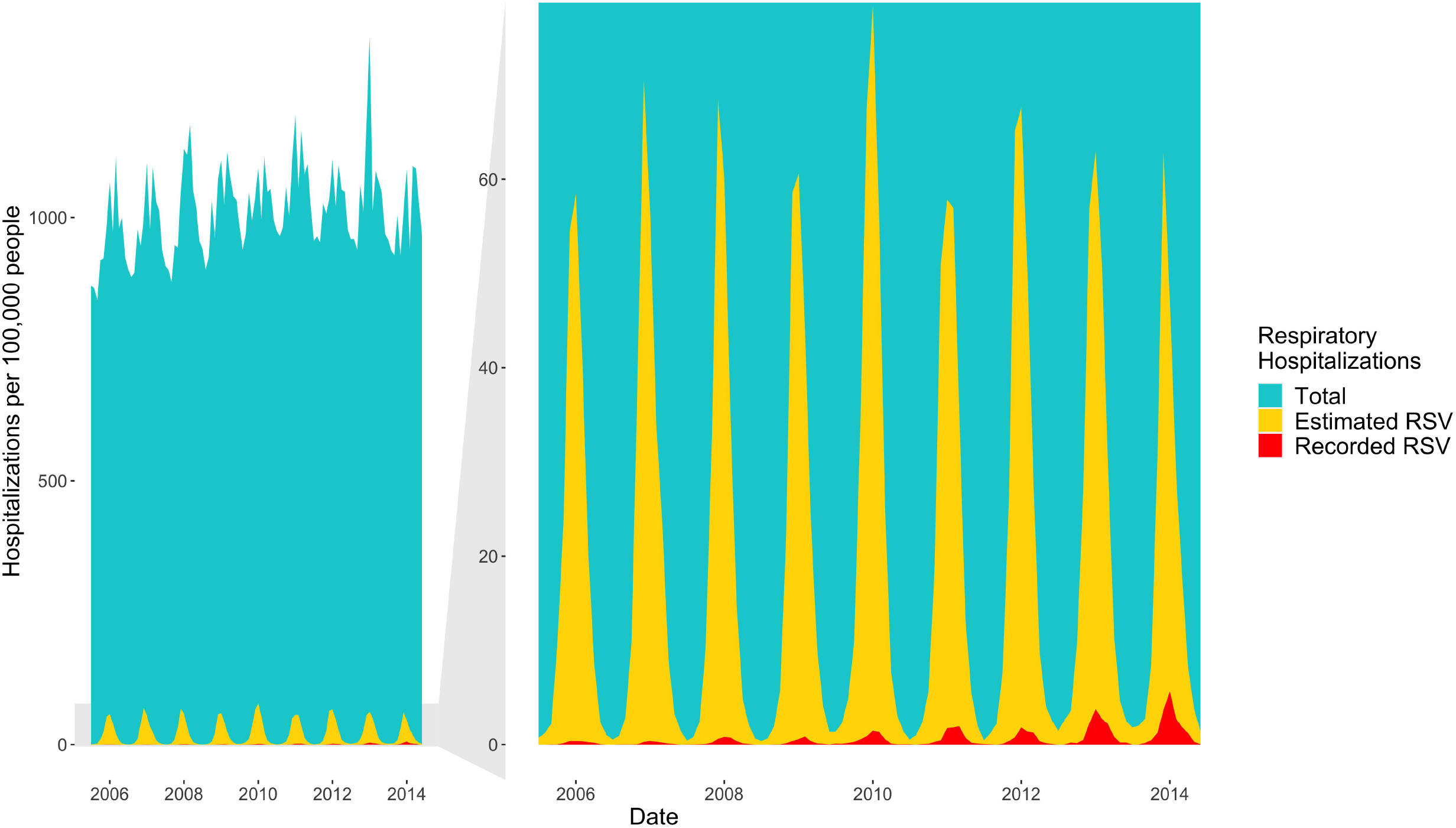
Respiratory hospitalizations among older adults, July 2005 - June 2014. The blue area in the left panel shows the total respiratory hospitalization rate among adults aged 65 and above; the yellow area shows the estimated RSV-associated respiratory hospitalizations in the same age group and the red area shows the hospitalizations recorded as RSV. The right panel zooms in to show the increasing trend of recording RSV diagnoses.

### Percent of respiratory hospitalizations attributable to RSV

Infections due to RSV were estimated to contribute to a large proportion of hospitalizations for respiratory illnesses in young children (Figure 4). On average, 45% (95% CrI: 42%-47%) of such hospitalizations were attributable to RSV infection in infants <1 year of age. This proportion was smaller in older age groups (Table 1). For example, in adults 20-44 years of age, only 1% (95% CrI: 0%-2%) of hospitalizations for respiratory infections were attributable to RSV infection. The attributable percent was similar across SES levels.

**Table 1.** Estimated Percentage of Respiratory Hospitalizations Attributable to RSV infections, by SES and Age Group, 2005–2014.

|  | Low SES |  | Medium SES |  | High SES |  |
| --- | --- | --- | --- | --- | --- | --- |
| Age (in years) | Mean | 95% CrI | Mean | 95% CrI | Mean | 95% CrI |
| <1 | 44% | (42%, 46%) | 46% | (44%, 48%) | 46% | (44%, 48%) |
| 1 | 27% | (24%, 29%) | 29% | (25%, 31%) | 27% | (24%, 29%) |
| 2-4 | 18% | (16%, 20%) | 21% | (18%, 23%) | 18% | (16%, 20%) |
| 5-9 | 9% | (7%, 11%) | 11% | (8%, 13%) | 9% | (7%, 11%) |
| 10-19 | 3% | (2%, 4%) | 4% | (2%, 5%) | 3% | (2%, 4%) |
| 20-44 | 1% | (0%, 2%) | 1% | (0%, 2%) | 1% | (0%, 2%) |
| 45-64 | 1% | (0%, 2%) | 1% | (0%, 2%) | 1% | (0%, 2%) |
| 65-84 | 2% | (1%, 3%) | 2% | (1%, 3%) | 1% | (1%, 2%) |
| 85+ | 4% | (3%, 6%) | 4% | (2%, 5%) | 2% | (1%, 3%) |

**Figure 4.**
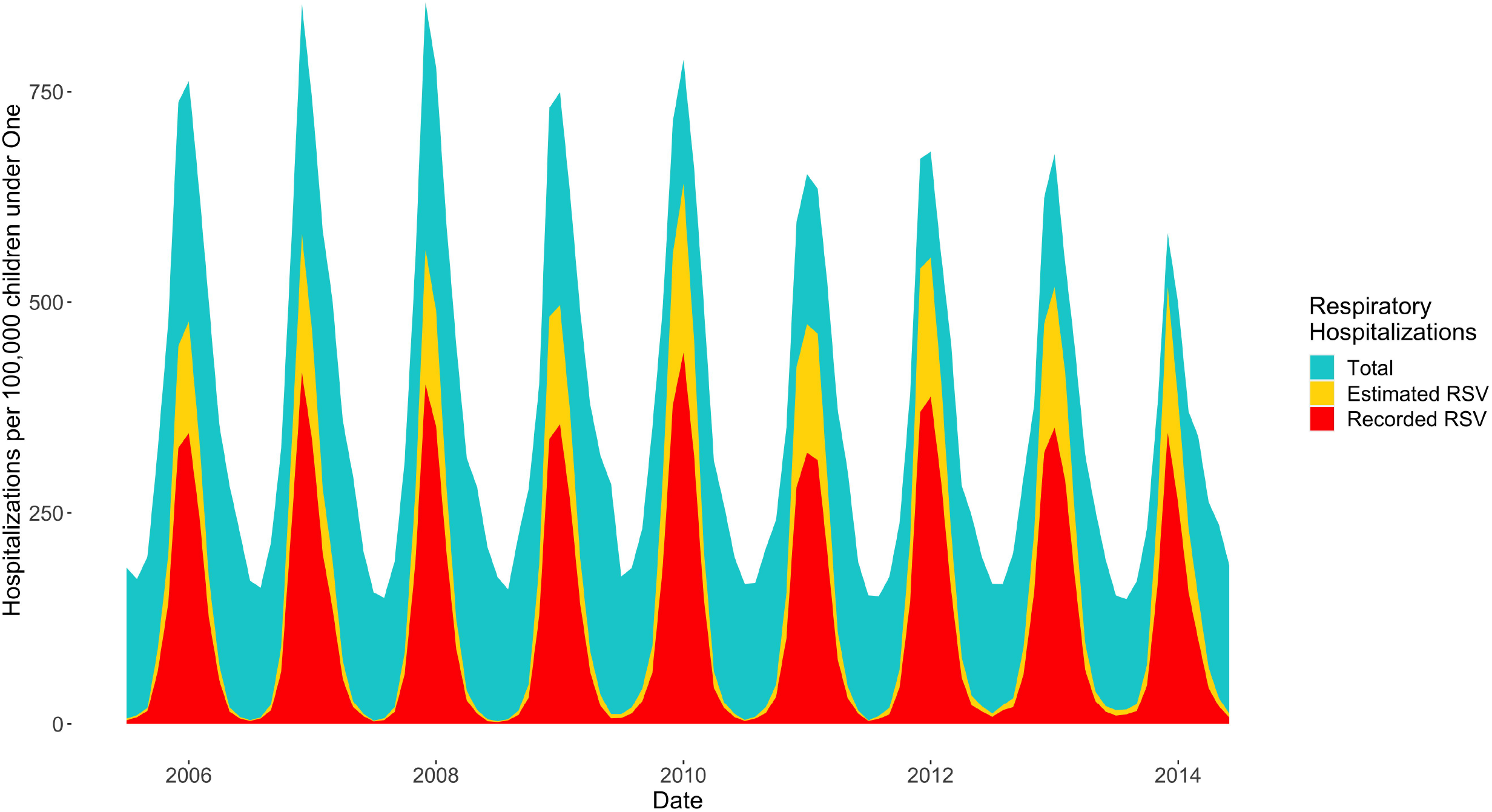
Respiratory hospitalizations in infants <1 year of age, July 2005 - June 2014. The blue area shows the total respiratory hospitalization rate in infants <1 year of age; the yellow area shows the estimated RSV-associated respiratory hospitalizations in the same age group and the red area shows the hospitalizations recorded as being due to RSV.

## Discussion

Understanding the incidence of hospitalizations caused by RSV can help to anticipate the potential impacts of upcoming RSV vaccine programs in different sub-populations. In addition, information about the differences in recording ratios between subpopulations may help increase awareness of providers about testing RSV in older adults and may lead to an improvement of RSV surveillance. We estimated a considerable burden of disease in young children (<5 years of age) and in the elderly (≥65 years of age). The estimated incidence of hospitalizations due to RSV was notably higher among patients from low SES ZIP codes. RSV was less likely to be diagnosed in older adults. The percentage of hospitalizations for respiratory illnesses attributable to RSV was highest in infants and lowest among adults 20-64 years of age. With several vaccines and monoclonal antibodies against RSV under active development, these estimates can help to guide estimates of the impact of these interventions in different populations.

Our age-based estimates for the incidence and attributable percent of hospitalizations due to RSV infections are consistent with previous studies ^18 27 28^. The statistical models we used in our analysis are an extension of the commonly employed time series models that estimate rates of respiratory virus-associated hospitalization ^15 29^. By modifying the model structure, our study provides new insights into the variation in RSV-associated respiratory hospitalizations by SES groups. Our estimates of the incidence of hospitalizations attributable to RSV infections among infants <1 year of age are lower than the average incidences in published estimates in the more distant past, but comparable to the averages in recent years ^18 27^.

Our estimates suggest that the incidence of hospitalizations caused by RSV has been under-recorded in the older adults. The substantial estimated incidence of hospitalizations attributable RSV among those aged 65 and older agrees with earlier observations in a cohort study and time-series study, both of which showed that the incidence of hospitalizations attributable RSV is heavily skewed toward the older adults ^30 27^. This age group should be considered as a potential target population for RSV vaccines due to the potential high case-fatality rate after RSV infection ^30 31^. The number of hospitalizations recorded as being caused by RSV increased over the study period in the patients 65 years of age and older. This trend may reflect changes in testing practices among older adults over time. To understand the actual incidence of hospitalizations attributable RSV in the elderly, more frequent testing for RSV infections is needed.

Our results indicate that children in low-SES communities suffer from a particularly high incidence of RSV-associated hospitalizations. There are a number of potential causes for this disparity, including factors that might influence risk of viral infection, such as family size and the number of contacts; exposure to tobacco smoke and other pollutants; high prevalence of underlying respiratory diseases like asthma and chronic lung disease from prematurity; and duration of breastfeeding ^32 33^. Additionally, decisions to for admit patients could be influenced by the family’s SES and may vary based on factors such as the co-morbid illnesses of the patients, the reliability of follow-up, and the practices of individual clinicians.

Our results suggest that about 45% of hospitalizations for respiratory illnesses in infants <1 year old are attributable to RSV infections (approximately 20 of 1000 infants/year). Monoclonal antibodies against RSV with an extended half-life ^34^, as well as vaccination of mothers and direct vaccination of infants using live-attenuated vaccines, might help to reduce the incidence of RSV infections ^31^.

There are several caveats to our results. First, we used hospitalizations due to RSV infections among children <2 years of age as a proxy for the timing of RSV infections in the entire population. However, there may be differences in the timing of infections among the various age groups. In our preliminary analyses, we tested different time lags between the various age groups, but it did not improve our model fit. Since our model used monthly inpatient data, minor differences in timing between age groups were less likely to be a factor. Second, the cocirculation of other respiratory viruses may confound our estimates. We may overestimate hospitalizations attributable to RSV by not including infections due to respiratory viruses other than influenza and RSV as covariates. However, previous studies indicated that most other respiratory viruses do not have the same timing as epidemics of RSV infections ^35-37^. Therefore, the cocirculation of other respiratory viruses should have a relatively small impact on our estimates of RSV-attributable hospitalizations. Third, the estimates of recording ratios rely on the validity of estimates of the incidence of RSV-attributable hospitalizations. Since there is no gold standard of estimates of RSV-attributable hospitalizations, it is hard to validate our estimates.

Still, our age-based estimates of recording ratios are similar to those in a previous study conducted by the Centers for Disease Control and Prevention ^27^.

In conclusion, children in families residing in low-SES areas had the highest incidence of RSV-associated hospitalizations for respiratory illnesses. The incidence of hospitalizations for RSV infections in the older adults is greatly under-recorded. More comprehensive testing for RSV among older adults might help to better define this problem. Vaccines against RSV might provide substantial benefit to young children and to the older adults.

## Data Availability

The demographic and geographic data that support the findings of this study are publicly available from the Geography program of the U.S. Census Bureau, the American Community Survey of the U.S. Census Bureau. The hospitalization data are not available publicly but can be obtained from the State Inpatient Database upon signing a data use agreement with the Agency for Healthcare Research and Quality. The R code for this study can be found in the GitHub repository: https://github.com/Weinbergerlab/RSVhospitalizations_USA/blob/main/RSV_burden.Rmd

## Author contributions

ZZ implemented the study, analyzed the data and drafted the article. JLW designed the study’s analytic strategy and helped prepare the Methods sections of the text. EDS helped revise the study design and manuscript for important intellectual content. VEP conceptualized the study and consulted on the analyses and revisions of the manuscript. DMW conceptualized and designed the study, and reviewed and revised the manuscript. All authors have seen and approved the final draft of the manuscript.

## Acknowledgments

The authors thank Dr. Louis Bont for constructive discussion that help the authors improve the manuscript.

## References

1. Hall CB, Weinberg GA, Iwane MK, et al. The burden of respiratory syncytial virus infection in young children. The New England journal of medicine 2009;360(6):588–98.

2. Shi T, McAllister DA, O’Brien KL, et al. Global, regional, and national disease burden estimates of acute lower respiratory infections due to respiratory syncytial virus in young children in 2015: a systematic review and modelling study. Lancet 2017;390(10098):946–58.

3. PATH. RSV Vaccine and mAb Snapshot. Secondary RSV Vaccine and mAb Snapshot April 2021. https://path.org/resources/rsv-vaccine-and-mab-snapshot/.

4. Single-Dose Nirsevimab for Prevention of RSV in Preterm Infants. The New England journal of medicine 2020;383(7):698.

5. Zheng Z, Pitzer VE, Warren JL, et al. Community factors associated with local RSV epidemic patterns: a spatiotemporal modeling study. medRxiv 2020:2020.07.06.20144345.

6. Noveroske DB, Warren JL, Pitzer VE, et al. Local variations in the timing of RSV epidemics. BMC infectious diseases 2016;16(1):674.

7. Simoes EA. Environmental and demographic risk factors for respiratory syncytial virus lower respiratory tract disease. The Journal of pediatrics 2003;143(5 Suppl):S118–26.

8. Homaira N, Mallitt KA, Oei JL, et al. Risk factors associated with RSV hospitalisation in the first 2 years of life, among different subgroups of children in NSW: a whole-of-population-based cohort study. BMJ open 2016;6(6):e011398.

9. Holmen JE, Kim L, Cikesh B, et al. Relationship between neighborhood census-tract level socioeconomic status and respiratory syncytial virus-associated hospitalizations in U.S. adults, 2015-2017. BMC infectious diseases 2021;21(1):293.

10. Lewis KM, De Stavola B, Hardelid P. Geospatial and seasonal variation of bronchiolitis in England: a cohort study using hospital episode statistics. Thorax 2020;75(3):262–68.

11. Green CA, Yeates D, Goldacre A, et al. Admission to hospital for bronchiolitis in England: trends over five decades, geographical variation and association with perinatal characteristics and subsequent asthma. Archives of disease in childhood 2016;101(2):140–6.

12. Cheung CR, Smith H, Thurland K, et al. Population variation in admission rates and duration of inpatient stay for bronchiolitis in England. Archives of disease in childhood 2013;98(1):57–9.

13. Lee N, Walsh EE, Sander I, et al. Delayed Diagnosis of Respiratory Syncytial Virus Infections in Hospitalized Adults: Individual Patient Data, Record Review Analysis and Physician Survey in the United States. The Journal of infectious diseases 2019;220(6):969–79.

14. Talbot HK, Falsey AR. The diagnosis of viral respiratory disease in older adults. Clinical infectious diseases: an official publication of the Infectious Diseases Society of America 2010;50(5):747–51.

15. Taylor S, Taylor RJ, Lustig RL, et al. Modelling estimates of the burden of respiratory syncytial virus infection in children in the UK. BMJ open 2016;6(6):e009337.

16. Simonsen L, Blackwelder WC, Reichert TA, et al. Estimating deaths due to influenza and respiratory syncytial virus. JAMA 2003;289(19):2499–500; author reply 500-2.

17. Zhou H, Thompson WW, Viboud CG, et al. Hospitalizations Associated With Influenza and Respiratory Syncytial Virus in the United States, 1993-2008. Clinical Infectious Diseases 2012;54(10):1427–36.

18. Goldstein E, Greene SK, Olson DR, et al. Estimating the hospitalization burden associated with influenza and respiratory syncytial virus in New York City, 2003-2011. Influenza and other respiratory viruses 2015;9(5):225–33.

19. Healthcare Cost and Utilization Project State Inpatient Databases. Agency for Healthcare Research and Quality. Secondary Healthcare Cost and Utilization Project State Inpatient Databases. Agency for Healthcare Research and Quality 2014. http://www.hcup-us.ahrq.gov/sidoverview.jsp.

20. American Community Survey 1-Year Data (2005-2019). United States Census Bureau. Secondary American Community Survey 1-Year Data (2005-2019). United States Census Bureau. https://www.census.gov/data/developers/data-sets/acs-1year.2010.html.

21. Casiano-Colón AE, Hulbert BB, Mayer TK, et al. Lack of sensitivity of rapid antigen tests for the diagnosis of respiratory syncytial virus infection in adults. Journal of Clinical Virology 2003;28(2):169–74.

22. Allen KE, Chommanard C, Haynes AK, et al. Respiratory syncytial virus testing capabilities and practices among National Respiratory and Enteric Virus Surveillance System laboratories, United States, 2016. Journal of clinical virology: the official publication of the Pan American Society for Clinical Virology 2018;107:48–51.

23. Bolker BM, Brooks ME, Clark CJ, et al. Generalized linear mixed models: a practical guide for ecology and evolution. Trends Ecol Evol 2009;24(3):127–35.

24. Gay NJ, Andrews NJ, Trotter CL, et al. Estimating deaths due to influenza and respiratory syncytial virus. JAMA 2003;289(19):2499; author reply 500-2.

25. Goldstein E, Viboud C, Charu V, et al. Improving the estimation of influenza-related mortality over a seasonal baseline. Epidemiology 2012;23(6):829–38.

26. Widmer K, Zhu Y, Williams JV, et al. Rates of hospitalizations for respiratory syncytial virus, human metapneumovirus, and influenza virus in older adults. The Journal of infectious diseases 2012;206(1):56–62.

27. Zhou H, Thompson WW, Viboud CG, et al. Hospitalizations associated with influenza and respiratory syncytial virus in the United States, 1993-2008. Clinical infectious diseases: an official publication of the Infectious Diseases Society of America 2012;54(10):1427–36.

28. Cassell K, Gacek P, Rabatsky-Ehr T, et al. Estimating the True Burden of Legionnaires’ Disease. American journal of epidemiology 2019;188(9):1686–94.

29. Fleming DM, Taylor RJ, Lustig RL, et al. Modelling estimates of the burden of Respiratory Syncytial virus infection in adults and the elderly in the United Kingdom. BMC infectious diseases 2015;15:443.

30. Lee N, Lui GC, Wong KT, et al. High morbidity and mortality in adults hospitalized for respiratory syncytial virus infections. Clinical infectious diseases: an official publication of the Infectious Diseases Society of America 2013;57(8):1069–77.

31. PATH. RSV Vaccine and mAb Snapshot. Secondary RSV Vaccine and mAb Snapshot. https://path.org/resources/rsv-vaccine-and-mab-snapshot/.

32. Li R, Darling N, Maurice E, et al. Breastfeeding rates in the United States by characteristics of the child, mother, or family: the 2002 National Immunization Survey. Pediatrics 2005;115(1):e31–7.

33. Prevention CfDCa. Cigarette Smoking and Tobacco Use Among People of Low Socioeconomic Status. Secondary Cigarette Smoking and Tobacco Use Among People of Low Socioeconomic Status November 25, 2019. https://www.cdc.gov/tobacco/disparities/low-ses/index.htm.

34. AstraZeneca. Nirsevimab MELODY Phase III trial met primary endpoint of reducing RSV lower respiratory tract infections in healthy infants. Secondary Nirsevimab MELODY Phase III trial met primary endpoint of reducing RSV lower respiratory tract infections in healthy infants.

35. Byington CL, Ampofo K, Stockmann C, et al. Community Surveillance of Respiratory Viruses Among Families in the Utah Better Identification of Germs-Longitudinal Viral Epidemiology (BIG-LoVE) Study. Clinical Infectious Diseases 2015;61(8):1217–24.

36. Price OH, Sullivan SG, Sutterby C, et al. Using routine testing data to understand circulation patterns of influenza A, respiratory syncytial virus and other respiratory viruses in Victoria, Australia. Epidemiology and infection 2019;147:e221.

37. Landes MB, Neil RB, McCool SS, et al. The frequency and seasonality of influenza and other respiratory viruses in Tennessee: two influenza seasons of surveillance data, 2010-2012. Influenza and other respiratory viruses 2013;7(6):1122–7.

