## Supplement for "Estimated Incidence of Respiratory Hospitalizations Attributable to RSV Infections across Age and Socioeconomic Groups"

**SUPPLEMENTARY TEXT**

**Supplementary Methods**

**Supplementary Results**

**Supplemental Methods:**

We compared three commonly used methods that adjust for seasonality and we found that monthly dummy variables resulted in significantly improved model fit compared to the other two methods (Table S1).

Table S1 DIC scores of competing seasonality components

| | Monthly Dummy | Polynomial Time Trends | Sinusoidal Curves |
|---|---|---|---|
| Deviance | 35954 | 38443 | 37671 |
| pD | 195 | 127 | 144 |
| DIC | 36149 | 38570 | 37815 |

The model structure of our hierarchical Bayesian regression is given as

$$Y_{ijk} \sim \text{Negative Binomial}(p_{ijk}, r), p_{ijk} = \frac{r}{r + \lambda_{ijk}}$$

where $Y_{ijk}$ denotes the number of all-cause respiratory hospitalizations at time $i$, in age group $j$, and SES group $k$; the expected value of $Y_{ijk}$ is $\lambda_{ijk}$; and the variance of $Y_{ijk}$ is $\lambda_{ijk}(1 + \lambda_{ijk}/r)$. The parameter $r > 0$ serves as an overdispersion parameter with $r \to \infty$ indicating that the mean and variance are the same, as in a typical Poisson regression framework. We define the expected value as a function of covariates and random effects such that

$$\lambda_{ijk} = \beta_{0jk} + \alpha_{1g(i)} + \alpha_{2m(i)} + \beta_{1jk}RSV_{ik} + \beta_{2g(i)jk}Flu_{ik} \quad (1)$$

where $\beta_{0jk}$ is the intercept parameter for age group *j* and SES group *k*; $\alpha_{1g(i)}$ represents an intercept term that varies by epidemiologic year, where *g(i)* is a function that maps time to epidemiologic year (defined from July in the previous year to June in the next year); $\alpha_{2m(i)}$ represents a similar intercept term which varies by month, where *m(i)* is a function that maps time to the corresponding month category; $\beta_{1jk}$ describes the group-specific effect of RSV, which associates RSV infections to respiratory hospitalizations; $\beta_{2g(i)jk}$ describes the association between influenza infections and respiratory hospitalizations, and varies by epidemiologic year in addition to the groups [27].

We model the coefficients of RSV infections ($\beta_{1jk}$ parameters) as a multiplicative combination of age and SES effects, such that

$$\beta_{1jk} = \exp\{\omega_{1j} + \gamma_{1k} + \epsilon_{1jk}\}$$

where $\omega_{1j}$ represents the age group effects; $\gamma_{1k}$ represents the SES effects; and $\epsilon_{1jk} \sim \mathrm{N}(0, \sigma_{\epsilon 1}^2)$ accounts for other unexplained variation. A similar structure is applied to the coefficients of influenza infections ($\beta_{2g(i)jk}$), with an additional yearly effect included, such that

$$\beta_{2g(i)jk} = \exp\{\omega_{2j} + \gamma_{2k} + \xi_{2g(i)} + \epsilon_{2g(i)jk}\}$$

where $\omega_{2j}$ represents the age group effects; $\gamma_{2k}$ represents the SES effects; $\xi_{2g(i)}$ represents the epidemiologic year effects potentially due to differences in the severity of the circulating strain; and $\epsilon_{2g(i)jk} \sim \mathrm{N}(0, \sigma_{\epsilon 2}^2)$ accounts for other unexplained variation. We assign weakly informative prior distributions to the remaining model parameters while ensuring that the remaining intercept parameters (i.e., $\beta_{0jk}, \alpha_{1g(i)}, \alpha_{2m(i)}$) are positive. Full details are given in the supplement.

Posterior samples were collected using a Markov chain Monte Carlo (MCMC) algorithm. To make posterior inference, three chains of 12,500 MCMC iterations were used following an initial burn-in of 62,500 iterations per chain. The combined set of 37,500 MCMC iterations were then thinned by a factor of 10, resulting in 3,750 less correlated posterior samples from the joint posterior distribution with which to make inference. Convergence was assessed by examining individual parameter trace plots and Gelman-Robin diagnostics [28, 29]. Posterior means and 95% equal-tailed quantile-based credible intervals were calculated using the samples. The model was fitted using the rjags package [30]

The hyperparameters for $\beta_{0jk}, \alpha_{1g(i)}, \alpha_{2m(i)}, \omega_{1j}, \omega_{2j}, \gamma_{1k}, \gamma_{2k}$ are as follows:

$$\beta_{0jk} = \exp\{\mu_{0jk}\}$$

$$\mu_{0jk} \sim \mathrm{N}(\mu_0, \sigma_0^2)$$

$$\alpha_{1g(i)} = \exp\{\mu_{1g(i)}\}$$

$$\mu_{1g(i)} \sim \mathrm{N}(0, \sigma_1^2)$$

$$\alpha_{2m(i)} = \exp\{\mu_{2m(i)}\}$$

$$\mu_{2m(i)} \sim \mathrm{N}(0, \sigma_2^2)$$

$$\omega_{1j} \sim \mathrm{N}\left(0, \sigma_{\omega_1}^2\right)$$

$$\omega_{2j} \sim \mathrm{N}\left(0, \sigma_{\omega_2}^2\right)$$

$$\gamma_{1k} \sim \mathrm{N}\left(0, \sigma_{\gamma_1}^2\right)$$

$$\gamma_{2k} \sim \mathrm{N}\left(0, \sigma_{\gamma_2}^2\right)$$

$$\sigma_0^2, \sigma_1^2, \sigma_2^2, \sigma_{\omega_1}^2, \sigma_{\omega_2}^2, \sigma_{\gamma_1}^2, \sigma_{\gamma_2}^2 \sim \text{Inverse Gamma}(0.01{,}0.01)$$

After we fitted the model, the estimated "true" number of hospitalizations attributable to RSV infections for each time point and stratum were estimated by multiplying posterior samples of $\beta_{1jk}$ by $RSV_{ik}$. The average annual incidence of RSV was estimated by dividing the sum of $\beta_{1jk}RSV_{ik}$ over nine epidemiologic years by the age- and site-specific population. The recording ratios were calculated by dividing the number of recorded ICD-9-CM diagnoses for RSV in each age and SES group by the modeled estimates in the same group. The attributable percent of RSV was calculated by dividing the sum of $\beta_{1jk}RSV_{ik}$ by the sum of the model-predicted number of all-cause respiratory hospitalizations in each age and SES group ($\lambda_{ijk}$) over the entire study period. (Note that $E\left(Y_{ijk}\right) = \lambda_{ijk}$ and $Y_{ijk}$ in each group are almost identical; $E(Y_{ijk})$ captures 99.98% of the variability in $Y_{ijk}$.) For each measure, we obtained and summarized samples from the posterior distributions of interest.

**Supplemental Results:**

Table S2 ICD-9-CM-coded respiratory hospitalizations in the entire populations and RSV hospitalizations among children <2 years in New Jersey, New York, and Washington.

| | ZIP | Respiratory hospitalizations | RSV hospitalizations |
|---|---|---|---|
| New Jersey | 581 | 2,555,728 | 170,703 |
| New York | 1715 | 5,482,926 | 328,833 |
| Washington | 512 | 1,379,736 | 100,575 |

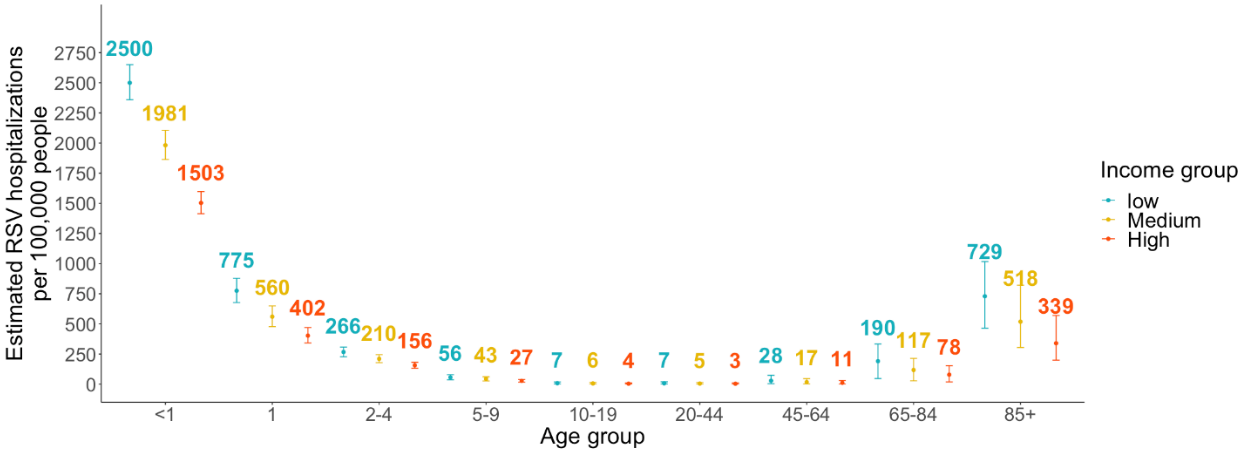


**Figure S1. Estimated RSV-attributable respiratory hospitalization rates by age and SES group in New York, July 2005 - June 2014.** The color texts show the mean estimates of RSV-attributable respiratory hospitalization rates in each age and SES group. The error bars indicate the 95% credible intervals of the estimated RSV-attributable respiratory hospitalization rates. Color blue, yellow, and orange correspond to the estimates in populations from low, medium, and high SES ZIP codes.

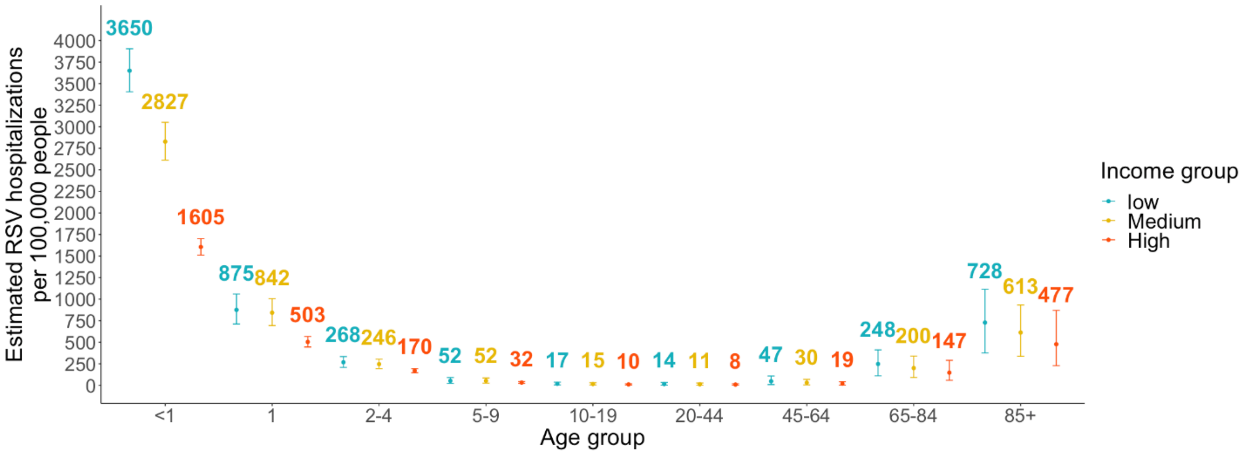


**Figure S2. Estimated RSV-attributable respiratory hospitalization rates by age and SES group in New Jersey, July 2005 - June 2014.** The color texts show the mean estimates of RSV-attributable respiratory hospitalization rates in each age and SES group. The error bars indicate the 95% credible intervals of the estimated RSV-attributable respiratory hospitalization rates. Color blue, yellow, and orange correspond to the estimates in populations from low, medium, and high SES ZIP codes.

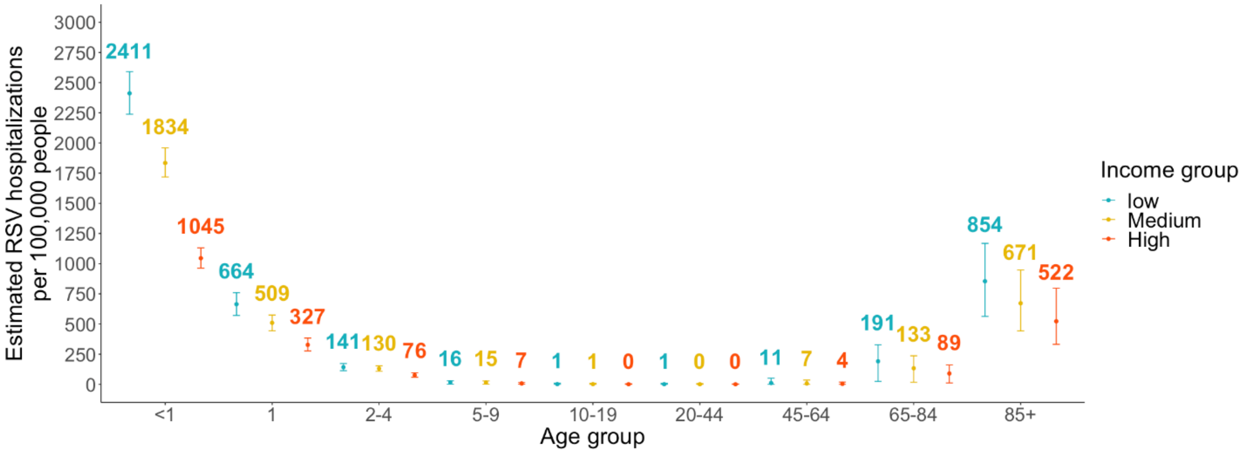

**Figure S3. Estimated RSV-attributable respiratory hospitalization rates by age and SES group in Washington, July 2005 - June 2014.** The color texts show the mean estimates of RSV-attributable respiratory hospitalization rates in each age and SES group. The error bars indicate the 95% credible intervals of the estimated RSV-attributable respiratory hospitalization rates. Color blue, yellow, and orange correspond to the estimates in populations from low, medium, and high SES ZIP codes.
